# Carers using assistive technology in dementia care: an explanatory sequential mixed methods study

**DOI:** 10.1101/2021.04.08.21255110

**Authors:** Vimal Sriram, Crispin Jenkinson, Michele Peters

## Abstract

**INTRODUCTION:** Informal carers support persons with dementia to live at home, even with deteriorating physical and cognitive issues. The purpose of this explanatory sequential mixed methods study is to examine the experiences and impact of Assistive Technology (AT) on carers, providing care for a person with dementia.

**METHODS:** The quantitative phase was a survey carried out with carers of persons with dementia in the UK, who used AT. The qualitative phase involved in-depth telephone interviews with a purposive sample of survey respondents. Data was analysed using hermeneutic phenomenology to develop, compare and explain the findings of the survey.

**RESULTS:** The survey included data from 201 carers. Smartphones and tablet computers were the most frequently used AT. Multiple AT were used in the care of persons with dementia predominantly for safety, communication, and reminders. The Short Form-12 questions in the survey showed that carers in the 46-65 age group and carers who were not extremely satisfied with AT had lower mental component scores and carers who lived with the person with dementia and older carers had lower physical component scores. Twenty-three carers participated in the interviews, and 5 themes with 14 sub-themes were identified. The interviews helped confirm data from the survey on the impact of AT on the physical, mental and social wellbeing of the carers. It helped describe reasons for satisfaction with AT; how AT was used in daily life and strengthened caring relationships; and how wider support systems enhanced the care of a person with dementia using AT.

**CONCLUSIONS:** This study describes the use of AT in the real-world context. AT supplements the care provided to a person with dementia in the community. Appropriate use, access to AT and abilities of the carer can enhance the support provided through AT to both carers and the person with dementia.

## INTRODUCTION

Dementia is a public health priority[1]. It is a progressive illness, and even if functional challenges impede a person’s ability to live independently, persons with dementia want to live at home[2]. Informal carers (family, friends and neighbours), hereafter referred to as carers, play a crucial role in supporting people living with dementia in the community. Assistive Technology (AT) is suggested as one way of providing support to the person with dementia and their carers[3,4]. AT can be defined as: “any item, piece of equipment, product or system that is used to increase, maintain or improve the functional capabilities and independence of people with cognitive, physical or communication difficulties”[5]. Carers usually make the decision on purchase, support maintenance and decide on abandonment of AT. While AT is viewed as a pervasive solution to supporting carers and persons with dementia to live for longer in the community[6–8], few attempts have been made to understand the experiences of carers, who use and support the use of AT. Understanding carers’ perceptions of AT and the impact of these ATs on carers is important to continue to provide support for persons with dementia and carers.

We carried out a survey among carers of persons with dementia in the UK[9]. This provided a broad understanding of the experiences of carers using AT. This survey provided information on the current use, satisfaction and impact of AT use among carers of persons with dementia. The survey was conducted using the Carers Assistive Technology Experience Questionnaire (CATEQ) [10] and the SF-12 (version 1)[11]. The SF-12 contains items covering physical functioning, social functioning, role functioning (physical and mental), vitality, bodily pain, mental health and general health. The SF-12 generates two summary scores: The Physical Component Score and the Mental Component Score (PCS and MCS respectively). The PCS and MCS are generated using norm-based methods and are standardised, using scores from the general population[11,12], to have a mean of 50 (SD 10). A higher score indicates better quality of life.

To gain a better insight into these findings, interpret them and to enrich the understanding of the experience and impact of using AT on carers, we used a sequential explanatory mixed-methods design[13–15]. In this method, quantitative data are collected and analysed first, then qualitative data are collected and analysed to help explain the quantitative data.

### Study objectives

This paper in addition to describing the results from the survey[9] provides context to the findings, by describing:

1. The experience of carers in using AT in the care of persons with dementia and
2. The impact of AT on carers well-being and daily life.

### Ethics and Patient and Public Involvement

This study was approved by the University of Oxford Central University Research Ethics Committee (Reference number: R57703/RE001). All potential participants were provided with a participant information sheet (supplementary file 1) and interview participants provided informed written consent (supplementary file 2) prior to the interview. No personal identifiable information of participants is reported in this paper. This study is part of a larger research project which has a patient and public advisory group that meets twice a year. This group consists of two carers of persons with dementia and a person with dementia (all living in England). This group reviewed the final version of the CATEQ and informed the interview guide questions. This group has also committed to support dissemination of study results to other patient involvement groups and their wider networks.

## METHODS

### 1.1 Study Design

This is a mixed-methods study [Fig 1], with the cross-sectional survey carried out between April to July 2020. Following the survey analysis, interview questions were formulated to gain further understanding into carers’ experiences as identified in the survey. Interview participants were recruited and interviewed between October – December 2020.

**Fig 1:**
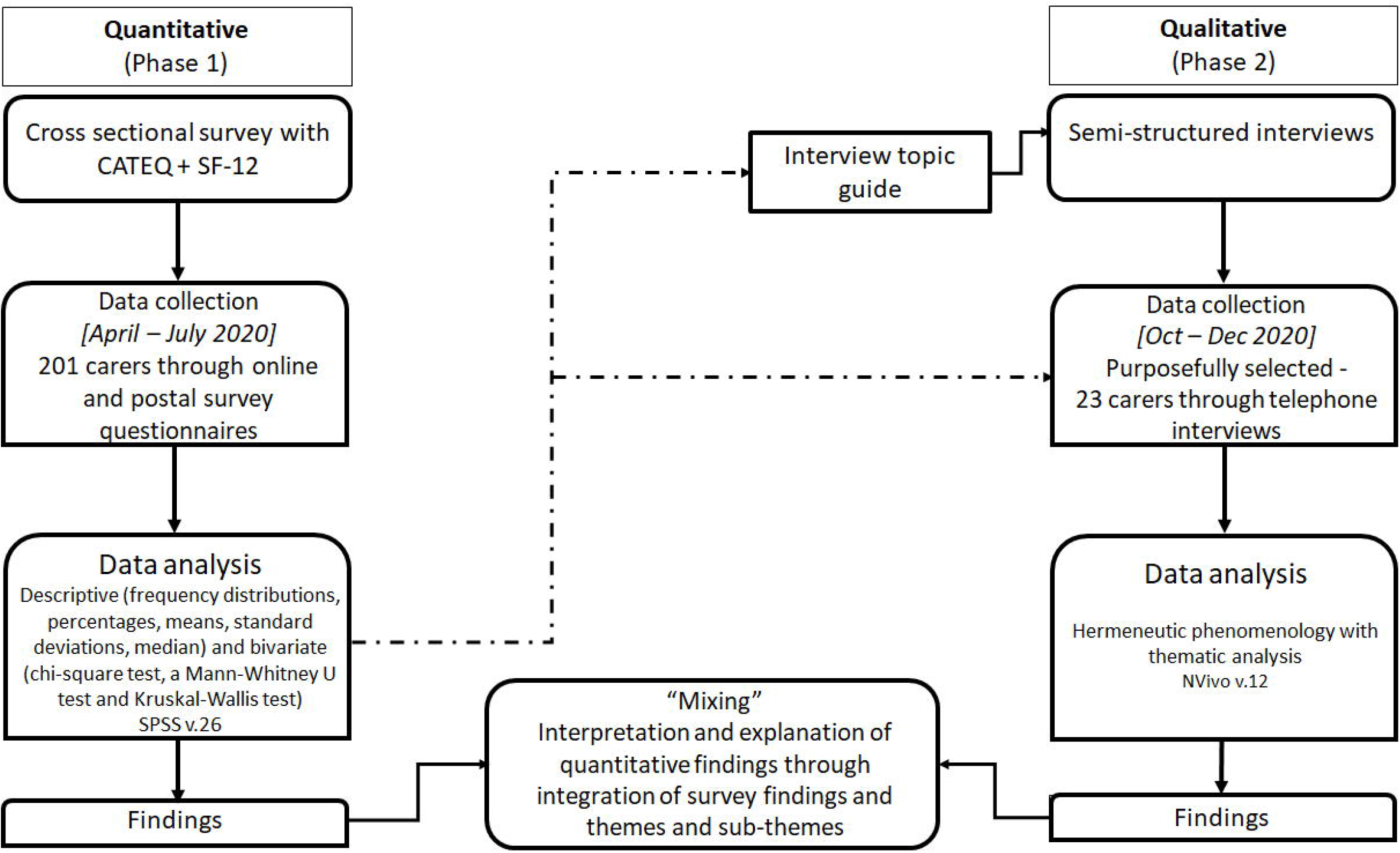
Flowchart of mixed methods design for carers using assistive technology in dementia care study.

### 1.2 Participants

#### 1.2.1 Recruitment

Survey participants were recruited using online databases via the Join Dementia Research website[16], Oxford Dementia and Aging Research (OxDARE)[17] and through health care professionals who prescribe AT for persons with dementia. Participants were carers of persons with dementia based in the United Kingdom. The inclusion criteria were: adult carers - family, friends or neighbours - providing at least 10 hours of care (e.g. shopping, leisure, personal care, finance) per week to a person with dementia who lives in their own home, with the carer living together with or away from the person with dementia; and carers needed to have used at least one AT device at home in the previous year. For the interviews, an email with the participant information sheet was sent to survey participants who gave consent to be contacted. From those who replied expressing an interest in participating, a purposive sample reflecting variations in age, sex, living arrangements, rurality, and relationship with persons with dementia were selected for the interviews.

#### 1.2.2 Data collection

For the survey, questionnaires were completed and returned by post or by using an anonymous online survey link. Data collection for the quantitative study is described in detail elsewhere[9]. For the qualitative study, semi-structured interviews were conducted over the telephone due to the restrictions from the COVID-19 pandemic. The interview focused on caregiving experience, the use and impact of AT on carers and on caring for the person with dementia. All questions and prompts were open ended and informed by an earlier systematic review[4], qualitative study[18] and the survey[9] and confirmed as meeting the needs for answering the research questions by the patient and public advisory group. All interviews were carried out by VS. Demographic data was taken from the survey responses and confirmed as being accurate with the participant at the start of the interview. The background of the interviewer as an Occupational Therapist and consequent interest in the research topic was discussed with participants. The participants were not known to the interviewer or the other authors before recruitment. Trust in the interviewer was built by establishing rapport with the participants through email and prior to answering questions as part of the telephone interview. Interviews lasted between 30-50 minutes, were audio-recorded, and later transcribed by a professional transcriber, verbatim with names of participants, names of the person with dementia, and any towns/cities mentioned in the interviews pseudonymised to ensure confidentiality.

### 1.3 Data analysis

#### 1.3.1 Quantitative data analysis

Survey data was analysed using IBM SPSS Statistics version 26. Descriptive statistics were used to examine the types of AT used, what the AT was used for, costs of the AT, perceived value for money and satisfaction with the AT. Bivariate analyses were conducted to examine differences in socio-demographic variables between respondents and SF-12 scores. The level of significance was set at p<0.05 for all analyses.

#### 1.3.2 Qualitative data analysis

The data was analysed using Hermeneutic Phenomenology, a process outlined by Lindseth and Norberg and others[19–21] and used in our previous qualitative study[18]. Hermeneutic phenomenology focusses on the subjective accounts of individuals own experiences in context, and thematic analysis seeks to identify and describe topic categories raised by interviewees in describing and explaining the survey results. VS listened to each of the interviews and read the transcripts multiple times (first step). Line by line coding of each transcript was carried out using the software package NVivo 12[22] (second step). The data analysis was ongoing throughout the period of data collection, this ongoing method allowed earlier transcripts to be recoded to reflect new codes. All the coded units were grouped into themes and underlying sub-themes (third step). Data collection continued after 20 interviews to ensure no new codes were developed and at the twenty third interview, it was concluded that data saturation was reached.

#### 1.3.3 Mixing analysis

The themes were developed, compared and interpreted in context of the survey results[23]. All authors coded selected transcripts independently and met regularly to discuss and agree ongoing data collection, generate themes, interpretation and integrating data analysis with survey results. Reflexivity (the examination of own beliefs, judgments and practices during the research process and how these may have influenced the research)[24,25] and integrity of the research process was maintained by all authors. The authors’ experience and previous research with people with long-term conditions, including dementia, provided the necessary expertise for this research. However, it is acknowledged that this experience may have influenced the coding and interpretation of the themes.

## RESULTS

Full results from the survey is reported elsewhere[9]. A brief overview of the findings is given here to provide context. Data from 201 carers was analysed. Participant characteristics from the survey is described in Table 1. AT were predominantly used for safety, communication, and reminders. Multiple AT devices were used in the care of persons with dementia and the number of AT used was associated with perceived satisfaction. Carers reported that AT helped them reduce effort of caring for a person with dementia and reduced perceived stress. Additional support was needed to support purchase and continued use of AT, and AT devices were abandoned when the person with dementia could no longer use them. A summary of the important findings from the survey is provided in Table 2.

**Table 1:** Characteristics of survey participants.

|  |  | N | % |
| --- | --- | --- | --- |
| Sex |  |  |  |
|  | Women | 131 | 65.2 |
|  | Men | 65 | 32.3 |
|  | Other | 1 | 0.5 |
| Living arrangements |  |  |  |
|  | Living with person with dementia | 103 | 51.2 |
|  | Living away from person with dementia | 98 | 48.8 |
| Ethnicity |  |  |  |
|  | White | 186 | 92.5 |
|  | Indian/Indian British | 4 | 2 |
|  | Mixed/multiple ethnic groups | 3 | 1.5 |
|  | Other | 1 | 0.5 |
| Marital status |  |  |  |
|  | Single | 17 | 8.5 |
|  | Married/civil partnership | 158 | 78.6 |
|  | Divorced/legally dissolved civil partnership | 22 | 10.9 |
|  | Widowed/surviving partner | 3 | 1.5 |
| Highest level of education |  |  |  |
|  | Secondary school | 8 | 4.0 |
|  | College (further education) | 58 | 28.9 |
|  | Undergraduate university degree | 76 | 37.8 |
|  | Postgraduate university degree | 51 | 25.4 |
|  | Other | 8 | 4.0 |
| Annual family income |  |  |  |
|  | Less than £10,000 | 7 | 3.5 |
|  | £10,001 - £40,000 | 86 | 42.7 |
|  | £40,001 - £70,000 | 49 | 24.4 |
|  | Greater than £70,000 | 11 | 5.5 |
|  | I do not wish to say | 47 | 23.4 |
| Relationship with person with dementia |  |  |  |
|  | Parent | 110 | 54.7 |
|  | Sibling | 3 | 1.5 |
|  | Friend | 2 | 1.0 |
|  | Neighbour | 1 | 0.5 |
|  | Spouse | 72 | 35.8 |
|  | Grandparent | 3 | 1.5 |
|  | Other | 10 | 5.0 |
| Age (Minimum – Maximum); Mean (SD) | 33 - 92 Years; 61.67 (12.07) |  |  |

**Table 2:**
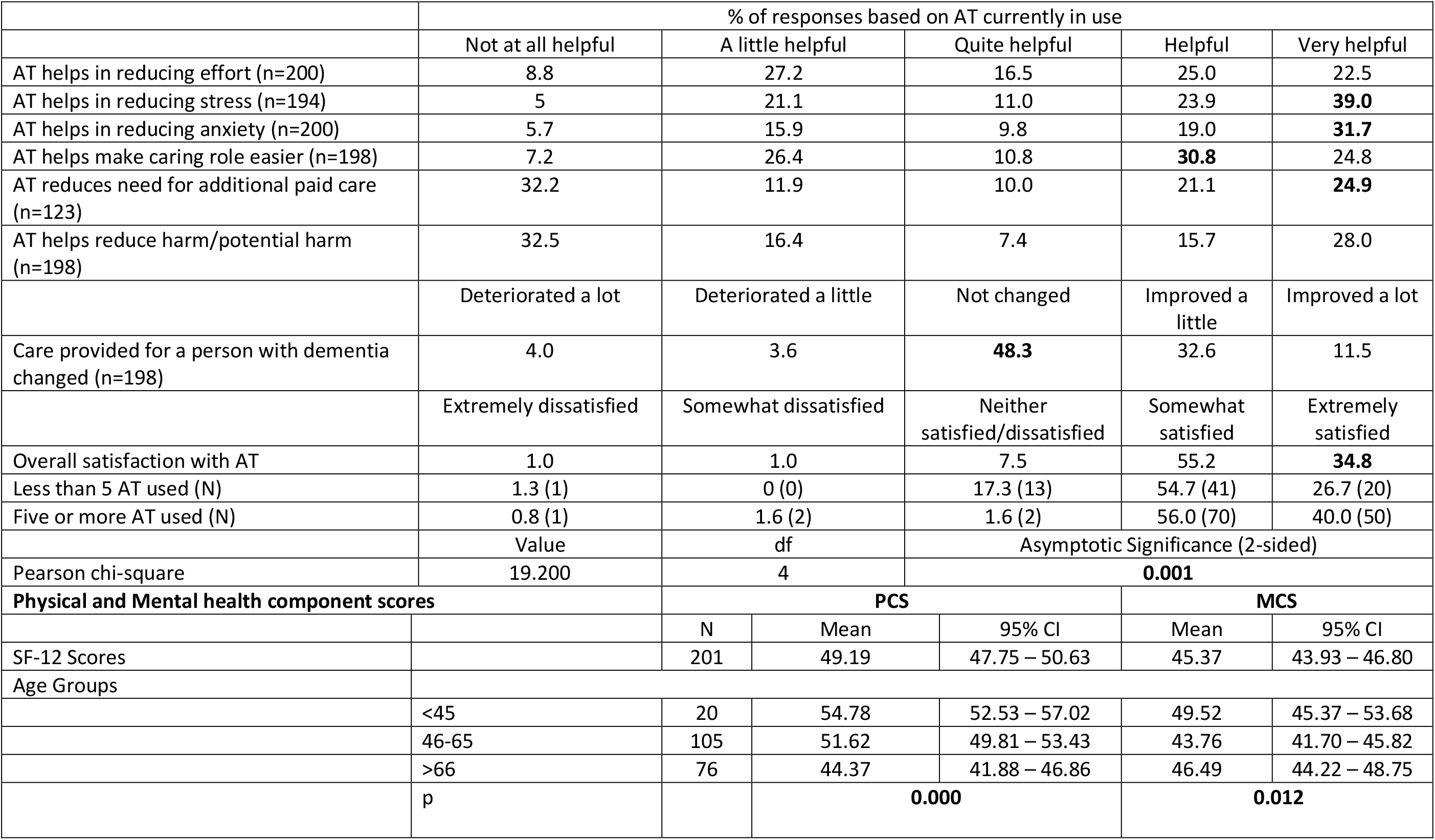
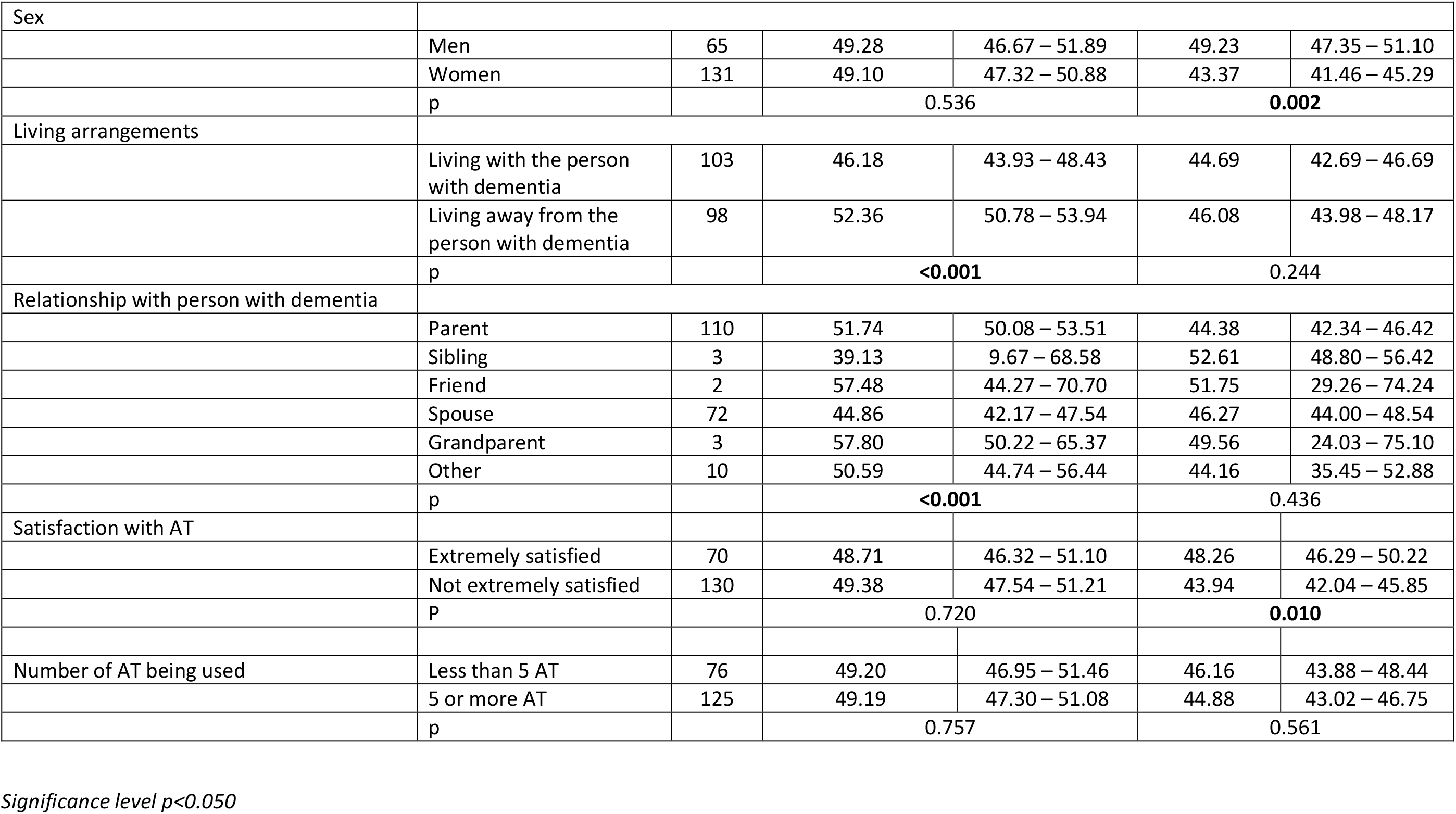
Findings from the survey: perceived impact of AT.

For the qualitative study 23 carers (18 women, 4 men, 1 non-binary) participated in the interviews. Participants’ age ranged from 51 to 85. Table 3 provides further details of the interview participants.

**Table 3:** Characteristics of interview participants.

| ID | Age Range | Gender | Person with dementia is | Ethnicity | Living arrangements | Assistive Technology used | PCS Score | MCS Score | Type of dementia | Years/months since diagnosis |
| --- | --- | --- | --- | --- | --- | --- | --- | --- | --- | --- |
| 1 | 71-80 | Female | Husband | White | Living with person with dementia | Laptop; cooker alarm; smart phone; stove timer | 23.11 | 52.03 | Vascular dementia | Two years |
| 2 | 51-60 | Male | Mother | White | Weekly visits | Audio books; automatic night lamp; dementia clock; GPS tracker; large button telephone; object locator; pendant alarm; picture button telephone; smart phone; web camera | 57.23 | 55.92 | Alzheimer's dementia | Eighteen months |
| 3 | 71-80 | Female | Husband | White | Living with person with dementia | Audio books; laptop; dementia clock; falls alarm; GPS tracker; pendant alarm; tablet computer; Alexa; web camera | 54.37 | 42.29 | Alzheimer's dementia | Three years |
| 4 | 51-60 | Female | Mother | White | Living with person with dementia | Automatic night lamp; laptop; smart gas meter; smart lights; tablet computer; web camera | 38.48 | 22.51 | Unsure | Ten years |
| 5 | 71-80 | Female | Husband | White | Living with person with dementia | Electronic reminders; large button telephone; smart phone; tablet computer; video communication | 39.90 | 35.17 | Alzheimer's dementia | Four years |
| 6 | 61-70 | Female | Mother | White | Living with person with dementia | Falls alarm; CCTV; GPS tracker; door alarm; memory clock; movement sensor; picture button telephone; web camera | 57.12 | 26.40 | Mixed dementia | Eight years |
| 7 | 81-90 | Male | Wife | White | Living with person with dementia | Laptop; dementia clock; community alarm; smartphone; video communications | 52.83 | 47.95 | Alzheimer's dementia | Three years, six months |
| 8 | 61-70 | Female | Mother | White | Living with person with dementia | Laptop; memory clock; tablet computer; video communications | 55.91 | 55.86 | Mixed dementia | One year |
| 9 | 61-70 | Female | Husband | White | Living with person with dementia | Automatic night lamp; baby monitor; laptop; cooker alarm; dementia clock; smart phone; stove timer; video communications; web camera; satnav in car | 53.95 | 47.56 | Mixed dementia | Six years |
| 10 | 61-70 | Female | Mother | White | Daily visits | Electric bed; dementia clock; falls alarm; GPS tracker; Large button telephone; simple radio | 62.47 | 28.33 | Alzheimer's dementia | Four years |
| 11 | 61-70 | Female | Husband | White | Living with person with dementia | Electric bed; memory clock; pendant alarm | 34.54 | 47.69 | Vascular dementia | Three years |
| 12 | 61-70 | Female | Mother | White | Visits every three weeks | Electric bed; stand aid | 50.91 | 50.12 | Unsure | Four years |
| 13 | 71-80 | Male | Wife | White | Living with person with dementia | Baby monitor; laptop; electric bed; smart gas meter; smart phone; video comms; Hoist; Wheelchair; WAV vehicle | 55.50 | 57.82 | Fronto-temporal dementia | Eleven years |
| 14 | 61-70 | Female | Mother | White | Daily visits | Laptop; dementia clock; GPS tracker; large button telephone; memory clock; pendant alarm; smart gas meter; smart phone; smart watch; video communications | 57.99 | 43.30 | Alzheimer's dementia | Seven years |
| 15 | 51-60 | Non-binary | Friend | Other | Daily visits | Laptop; electronic reminders; large button telephone; video communications; web camera | 56.44 | 49.98 | Parkinson's Dementia | One year, three months |
| 16 | 61-70 | Female | Husband | White | Living with person with dementia | Assistive robot; automatic night lamp; laptop; electronic reminders; GPS tracker; smart phone; tablet computer; video communications; Alexa | 24.31 | 35.38 | Alzheimer's dementia | Four years |
| 17 | 71-80 | Female | Husband | White | Living with person with dementia | Laptop; cooker alarm; falls alarm; pendant alarm; smart gas meter; smart phone; smart plugs; stove timer; tablet computer; video communications; Alexa; web camera | 33.30 | 27.00 | Vascular dementia | Twelve years |
| 18 | 71-80 | Female | Husband | White | Living with person with dementia | Pendant alarm | 44.47 | 38.91 | Lewy body dementia | Seven years |
| 19 | 51-60 | Female | Mother | White | Living with person with dementia | Audio book; laptop; dementia clock; GPS tracker; object locator; smart gas meter; smart phone; tablet computer | 62.85 | 26.05 | Alzheimer's dementia | Two years |
| 20 | 71-80 | Male | Wife + Mother-in-law | White | Living with person with dementia (wife). Mother-in-law recently moved to nursing home | Cooker alarm; dementia clock; GPS tracker; smart gas meter; smart lights; smartphone; tablet computer; video communications; ELK lifting cushion | 51.21 | 33.58 | Behaviour variant fronto-temporal dementia (wife) + Vascular dementia (mother-in-law). | Four years (wife)<br>Unsure (mother-in-law) |
| 21 | 51-60 | Female | Mother + stepdad | White | Weekly visits (mother recently | Falls alarm; Large button phone; memory clock; pendant alarm; | 60.39 | 27.12 | Alzheimer's dementia (mother) + | Five years |
|  |  |  |  |  | moved to nursing home) | picture button telephone; Video communications |  |  | Vascular dementia (stepdad) |  |
| 22 | 51-60 | Female | Father | White | Daily visits | Dementia clock; door alarm; tracking device; GPS tracker; memory clock; smart phone | 49.85 | 43.63 | Alzheimer's dementia | Four years, six months |
| 23 | 51-60 | Female | Father | White | Daily visits | Electric bed; smart lights; tablet comp; video communications; Alexa; movement detector | 50.88 | 17.41 | Mixed dementia | Two years |

### Themes

Five themes and 14 sub-themes were identified in the analysis. We describe the themes from the qualitative interviews and situate them in context of the survey results. Additional illustrative participant quotes are provided in Table 4.

**Table 4:** Themes and sub-themes with illustrative quotes.

| Theme | Sub-theme | Example quote 1 | Example quote 2 | Example quote 3 |
| --- | --- | --- | --- | --- |
| Use of AT | Staggered purchase and use of AT | we got things as, as she worsened [Participant 2] | Well as we got problems we found these technological solutions to enable us to continue to care for him at home [Participant 23] | Yeah. I, I got them, I think, gradually as Mum's condition progressed ... I was trying to maintain her independence as long as possible [Participant 6]. |
|  | Ease of using AT | I think it's [electric bed], it's much more help than, than anything else, no. I mean, we, we couldn't ... As I say, because she still can cooperate with using it [Participant 12] | I mean, I am very, I'm, I'm not au fait, au fait with it all but what I do know I'm, I'm able to use quite efficiently [Participant 4] | His own phone, he's okay answering it and doing the odd text with one word. I, I, I've got to admit, though, his iPad has been beneficial to him [Participant 5] |
|  | Problems using AT | ... but she would take it [pendant alarm] off from her neck and just throw it onto the settee which activated the alarm [Participant 14] | But there was, there was at one point the, the hoist that, it hadn't charged and so he got stuck half-way [Participant 18] | ...GPS tracker on her phone but after a while she forgot to, to take her phone with her, so when she went walkabout we had no idea where she was, and that was something that was really problematic [Participant 2] |
| Satisfaction with AT | Ability of the PwD | she's got rheumatoid arthritis [Participant 15] | She's got quite severe depression... She won't come downstairs, she refuses, she won't go out in the garden. She's in that room and that is it [Participant 4] | She can't remedy any mistakes that she makes. She gets very frustrated, she panics and then she presses all sorts of buttons and then calls us [Participant 14] |
|  | Problem solving | I've got a little key finder, which I call my mum finder, and I always take that out with us as well and slip it in her pocket or something, and again it's on a lanyard so she will play with it, and it's just more if we get separated [Participant 19] | So what we did instead was use the pad, these pressure pads that you put under the seat, under the cushion. So as he started to get up out of the chair, it would, it's wireless...the beep would go off and I could go and make sure he was okay. [Participant 23] | Well, I can, I can use it, but I don't find any technology particularly easy. I've always got to sit and think through it, you know? [Participant 10] |
|  | Strengthened relationships | That [CCTV camera] has helped because, and obviously being able to see him, helps him because he thinks, yes, there's people out there that care about him [Participant 23] | I think it's [AT], it's maintained a very close and stable relationship that was always there. It's, it's just experienced in a different way [Participant 7] | It [Youtube on tablet computer] has helped me and mum because instead of just sitting there keeping her occupied, and doing puzzles and, and chatting and watching, we've been able to do the tai chi [Participant 10] |
| Impact of AT on carers | Physical wellbeing | It affected me physically because I ended up having to either lift him up or help him into bed or get him into bed, or get him into a wheelchair, get him into the car [Participant 18] | I do a little workout routine with Mum in the morning to YouTube [Participant 19] | For my quality of life, but physically, without that [electric bed and hoist] assistance, I could not have managed [Participant 13] |
|  | Mental wellbeing | My dad was always the one that we went to when we needed to speak to someone, and that support was gone [Participant 22] | I can't, I can't begin to tell you what, you know, how much, how much, how helpful they [GPS tracker, movement sensor] were and how much anxiety they took away. [Participant 6] | I tend to use the technology for my own amusement at times [Participant 20] |
|  | Social wellbeing | It [Smart phone and tablet computer], it's absolutely helped to sustain my social life [Participant 17] | Yes, it's helped there [socialising] definitely...it does mean that one person can look after my dad at a time, rather than two people being there if you need to do lots of things [Participant 23] | We can communicate with our daughter and grandson in [city] via WhatsApp or anything and actually see them while we're talking to them which has obviously made a difference if she does that [Participant 7] |
| AT use in daily life | Coping with caring | Yes, we wouldn't be able to function at all without those[Electric bed; memory clock; pendant alarm] [Participant 11] | You know, when you have worked out routes for him to take, the length of time it takes him to walk [using the GPS tracker] and basically if he not back within that time then I, then I would have to go and see what's happening [Participant 16] | I could go out more often, and maybe not going more than a quarter of an hour from home or something like that, so I could get back if a problem arose [find out using the CCTV camera, smartphone] or, or I could ring the next door neighbours and say, 'Please go and sort her out' [Participant 20] |
|  | Person with dementia using AT | He watches television which is good for stimulating him but he can't use the remote control, so he relies on me to sort of get it all set up for him [Participant 11] | When she comes over to us in the evening then we will, do a Zoom or we'll do a, a Facetime or, a WhatsApp with our children so that she can see them, but left to her own devices she couldn't do it [Participant 14] | An internet device which calls my phone if she has a fall or if she wants to get in touch and she can talk to me over it. It's [community alarm] becoming less and less useful, because she can't do anything ... she can't go anywhere on her own [Participant 7] |
|  | Simple devices | We have a visual calendar in his, in his kitchen that we write things on that are happening and he really likes that [Participant 21] | So my idea is to keep regular photos of the family where possible so that she's aware of how they are changing and who they are [Participant 8] | The key safe outside, it means if at all necessary, if she forgot. Well, she wouldn't know how to use it, but we would always have a key if, if we had to run up in an emergency or something, there would be a key there [Participant 10] |
| Wider support systems | Support from others | it's [formal carer visits] a, it's a safety net and it also enables mum to see somebody else [Participant 14] | I now have a private carer who comes to help me one hour in the morning Mondays to Fridays and one hour in the evening Tuesday, Wednesday, Thursday just to give me a hand [Participant 9] | Even though there is a carer there as well moving her and doing a lot of things needs two people now [Participant 12] |
|  | Ethical issues | The [GPS] tracker, she doesn't know what it is. I sneak it on her with her sunflower lanyard when we have to go in shops [Participant 19] | We didn't tell him what it was for, we just said that there was a button on it that he could press if he needed us and it would ring on the phone here. But we didn't actually tell him that we knew where he was going or that we could see where he was because he wouldn't have accepted it [Participant 22] | I think personal security, personal secrecy worries me a little bit on that front [Participant 20] |

### 1. Use of AT

In the survey, carers indicated using a wide variety of AT including smart phones, tablet computers and video monitoring systems. They also reported staggered purchase or use of AT. Some AT devices such as pendant alarms and audio books were frequently abandoned as the person with dementia was no longer able to use them.

#### a. Staggered purchase and use of AT

Interview participants confirmed that as a person with dementia’s needs worsened, AT devices were added to support them and as these AT were purchased to meet specific needs, the satisfaction with these devices were higher.

> “the clock was one of the first things that I got because she really struggled with time and day”. [Participant 19]

Computers and tablet computers were already in use by carers or the person with dementia and were adapted to support caring needs

> “Well, I already had the laptop…and she started using it for memory [games]”. [Participant 13]

Carers had to anticipate future needs of the person with dementia and added the AT devices as and when a need arose.

> “The increase in [motion sensor] alarms has gone on as he has deteriorated and that is really because of the danger of him falling downstairs…it’s had to be a gradual, a gradual putting-in and I’ve still, you know, I’ve still got one or two things, one or two plugs that I need to change on the Hive [smart home technology] system.” [Participant 17]

Some of the AT procurement was constrained due to the carers’ own knowledge of what AT was available and what devices could support the person with dementia.

> “I think in all honesty I’d have liked to have had that one [CCTV cameras] before but didn’t really know what would be suitable and again, lacked the technological know-how to say how do I sort this out.” [Participant 9]

#### b. Ease of using AT

The ease of using AT had an impact on satisfaction, continued use or abandonment of AT. This in turn affected the impact the AT had on carers.

Carers found some AT helped a person with dementia, even when they lived away. This provided the carer with a sense of reassurance.

> “The tile [object locator] certainly helped so that remotely we could make them [objects] buzz for her and that made it easier to find things”. [Participant 2]

However, carers also reported that persons with dementia struggled to use AT such as pendant alarms, as intended

> “she’s supposed to press [pendant alarm] if she falls? But she, she can’t remember what it’s there for. She puts it on religiously, it’s like putting on jewellery. But the twice she has fallen in the house, she’s never pressed it”. [Participant 10]

Elderly carers who were not used to technology and digital devices, initially struggled with AT and described the need for access to support and education.

> “I think I’ve done extremely well with it [AT] but I’ve possibly … It, it would have been useful to have some access to education about that”. [Participant 9]

#### c. Problems using AT

While most of the AT devices that were currently in use were perceived as being user-friendly, carers also reported problems in the AT themselves or in their design. Some of the AT equipment such as electric beds and electric hoists were large and unwieldy in a home environment.

> “I mean, all the equipment’s large, but that [electric hoist] is really cumbersome”. [Participant 18]

Some of the problems encountered with AT was a result of the ability of the person with dementia to look after the devices.

> “Now the problem with the electronic alarm, the electronical alarm watch lasted about four months and then it got dropped in the bath by [person with dementia]”. [Participant 15]

The design features of some of the AT also caused problems, did not serve their intended purpose or caused confusion in the person with dementia.

> “The GPS [tracker], that is a right pain because it has to be charged up every night”. [Participant 20]

> “But, again, because, you know, you’ve got to have the angles and, and everything right and sitting comfortably, that [video phone call] works up to an extent, but I think after a while when we’ve, we’ve done the [video] call she’s sort of asking, “Well …” you know, “… where are they?” She can’t sort of understand the concept of what’s going on”. [Participant 8]

Carers wanted AT developers to involve them as end users, to ensure devices are easy to use and are accessible to those who are technologically challenged.

> “perhaps if they just sought a bit of feedback, we could help them just alter things a little bit, you know, not, not greatly, just to make things even more accessible for people and, and for us”. [Participant 10]

### 2. Satisfaction with AT

The survey showed that carers who had more than 5 AT devices expressed more satisfaction from using AT. During the interview, carers confirmed how they made the decision to use AT to support a person with dementia based on their needs, which included other co-morbidities not associated with dementia and described how the use of AT strengthened their relationship.

#### a. Ability of the person with dementia

While most of the AT were used to support difficulties a person experienced, because of dementia, some of the AT were used to support a person with dementia for other illnesses.

> “We use it [electric bed] as unfortunately she’s also got severe osteoarthritis”. [Participant 12].

Persons with dementia benefitted from AT to support them with cognitive issues, however as they were still physically active, carers continued to struggle with providing adequate care. Some AT were later put in place when the situation of the person with dementia deteriorated.

> “The first years [since diagnosis] were in some ways more difficult because she was still active and would go walkabout… She then broke her hip and that was it, her mobility went. She wouldn’t bear weight thereafter…so we had to use these equipment [electric bed and hoist]”. [Participant 13]

Carers also used AT to support persons with dementia, when their own abilities meant they could no longer participate in activities with the person with dementia.

> “So, he quite likes walking and getting about and I can’t no longer do the length of walk that he would like so we use the tracker [GPS tracker device]”. [Participant 3]

Some of the AT were also abandoned when the person with dementia’s situation worsened due to non-dementia related reasons.

> “He used, he really lost the ability to use a computer [cognitive reasons], but he could, he did until maybe 18 months ago [now no longer using], cause his eyesight really was getting worse, he used a Kindle [eBook reader]”. [Participant 18]

#### b. Problem solving

Carers described how their ability to problem solve when the AT failed assisted them in the continued use of the technology.

> “So, she has one friend where he phones her up and he painstakingly talks her through how to get into Skype on the computer…then he can hang up and they can have a conversation ‘cause he lives in Germany’”. [Participant 15]

Carers also simplified the use of some off-the shelf devices to suit the specific needs of the person with dementia.

> “He’s got some, a couple of photographs and my number, and my daughter’s number [on the smart phone]. And we had other things on it but we’ve had to limit it to that [two numbers] because he can manage to, to press and ring with those two [being] very visible, but that’s now what we’ve had to reduce it down to”. [Participant 17]

#### c. Strengthened relationships

While some survey participants described that AT gave additional time for themselves and the use of AT did not make a difference to their relationship with the person with dementia, most carers described that the AT helped strengthen their relationship with the person with dementia. This was confirmed by interview participants as another reason for their increased satisfaction with using AT.

Carers felt that the AT helped provide reassurance to the person with dementia, that the carer was still trying to keep them safe.

> “I suppose he [person with dementia], because it’s, he’s not able to communicate that well now, but hopefully it helps him to realise that I am coping and keeping him safe with the equipment that we use”. [Participant 11]

AT also helped some carers spend more time together with the person with dementia

> “We can sit down and watch something together [on tablet computer] and, and engage with it, and, you know…and stuff and we can have a giggle and then you end up talking about old neighbours or old family members. So, yes, it does help”. [Participant 19]

### 3. Impact of AT on carers

Survey participants in the 46-65 age group and carers who were not extremely satisfied with AT had lower mental health component scores, carers who lived with the person with dementia and older carers had lower physical component scores on the SF-12. The interviews explored if AT had an impact on carers and to explain the variations in SF-12 physical and mental health component scores.

#### a. Physical wellbeing

Carers had multiple physical health problems, this was not always a result of caring or looking after a person with dementia, age or illness associated physical health problems. They reflected on their own health, which may explain the lower scores especially among older carers.

> “As for myself, I haven’t got very good health. I’m suffering from very bad back pain”. [Participant 1]

While some of these issues were long standing, a few physical health issues arose because of caring for the person with dementia.

> “I mean, I have picked him up and ended up in hospital because I had done the disc in my back”. [Participant 17]

Carers also made changes to their lifestyle as part of the caring role for the person with dementia.

> “Even things like, you know, my diet is affected, because he has to be fed completely and his ability to chew and swallow is, is impaired. So, therefore I tend to think, “Oh, well, I’ll eat just whatever he’s eating”. [Participant 9]

In this context, AT did have a positive impact on the physical wellbeing of carers, especially for moving and handling of the person with dementia.

> “It’s certainly taken the strain off of my body, off my muscles and off my back. So it’s been a great help, both the [electric] bed and the hoist”. [Participant 11]

The reassurance provided by AT used for safety, especially for carers who lived away from the person with dementia, assisted carers in better sleep and maintaining an active and healthy lifestyle.

> “So I can fall asleep better [carer using door alarm for wandering alerts]”. [Participant 22]

#### b. Mental wellbeing

Caring for a person with dementia did cause a strain on the mental health of carers.

> “so it created a huge amount of stress as Mum was deteriorating being on her own all the time”. [Participant 19]

Some of the issues were indeed compounded by carers having to look after their own families in addition to a parent.

> “my son, he has Asperger’s, so at home it can be quite stressful at home at times”. [Participant 21]

The use of AT did offer relief to carers and provided reassurance to them.

> “So, it means we know that she’s taken her tablets. We know that there’s music if she wants to. We know she can use the bed”. [Participant 10]

Some of the AT also filled a need amongst carers for their own leisure and communication needs.

> “I would be lost without my technology, socially, mentally I need it…I still run a [virtual] group but I also am able to join other groups”. [Participant 17]

The AT also provided much needed time to carers for themselves and do tasks that they wanted to complete.

> “Well, I think it’s just that I can entertain Mum [using tablet computer] so that I can have some time to just catch up on paperwork or make a phone call”. [Participant 19]

#### c. Social wellbeing

In addition to physical and mental health impacts, AT also had an impact on social wellbeing of carers and the person with dementia.

> “So, having the Facetime or the WhatsApp [on smart phone] has been very good, so she can see them [family] and keep, keep in touch”. [Participant 14]

It also provided an opportunity for carers to continue to engage in social activities that they enjoyed before the diagnosis and progression of illness for the person with dementia.

> “we were still able to use assistive technology [GPS tracker, web camera] while mobile so that we could, we could still go out and about and be able to get the alarms or the, the contact if there was an issue [Participant 2].

### 4. AT use in daily life

From the survey, carers revealed that the AT had not significantly changed the level of care provided to the person with dementia by them but reported that AT helped in reducing the need for paid/formal care, which was one of the reasons for carers recommending AT use to others.

#### a. Coping with caring

AT is perceived by carers as supplementary to the care that they provide. As the abilities of the person with dementia deteriorates with progress of the illness, AT helps carers cope with caring needs.

> “We would look at the cameras if we were uncertain whether she had eaten fully”. [Participant 2]

AT was perceived as being integrated into the daily life and routines of the carers, especially when it concerned the safety and welfare of the person with dementia.

> “Well, all of them [CCTV, movement sensor, door alarm] become integrated. I mean I’ve got the front, the exit monitors on all the time… because, you know, you can’t be in the same room as them [person with dementia] all the time, can you?”. [Participant 6]

Some carers also felt that the use of AT increased the frequency of checking-up on the person with dementia and added to their care providing tasks.

> “So, we’ve just resorted to checking her frequently [using CCTV], having the carer check her once a day … that’s the best thing that we can do … Oh, it’s, well, the monitoring of it has taken over our life”. [Participant 14]

#### b. Person with dementia using AT

Carers also reported that persons with dementia had varied ability in using the AT by themselves, which meant carers had to continue to support them in the use of the AT.

> “Despite the fact it has the day and the date [dementia clock] she [Person with dementia] still sits and asks what day it is … so, we say, ‘Well, have a look at the clock’… the radio, I thought it would be more useful than it is. It’s all, it’s quite easy to set up, but I think with all of these things, they all require lots of support from somebody else”. [Participant 10]

Persons with dementia continued to struggle with using AT even with prompts from carers.

> “When the alarm goes off, she goes, ‘What’s that, what’s that noise?’ ‘It’s your alarm watch, are you gonna switch it off?’… ‘No, no’ and she just wouldn’t switch it off, she wouldn’t do anything with it” [Participant 15]

#### c. Simple devices

Even though carers used AT, they combined these with non-electronic or simple devices/solutions that worked in supporting and caring for the person with dementia. This combined use of simple and electronic AT proved more effective in the level of care provided as well as reducing the stress and anxiety associated with caring.

> “I also write our shopping lists on there [white board] so, anything we’ve run out of I write on there, or [Person with dementia] writes on there. So, we, we write our shopping list out and then I’ll wipe it all off afterwards. So, it is very, very handy”. [Participant 1]

Carers also adapted existing devices when the intended use of the AT did not support the person with dementia.

> “And I’d adapted a seven-day pill box to make, you know, to, to make sure there’s, that each compartment is one particular time [adapting electronic pill dispenser]”. [Participant 15]

### 5. Wider support systems

In the survey, carers reported that AT is an adjunct to supporting a person with dementia. The costs, privacy concerns, support from wider care networks and support systems all played a role on how carers used AT.

#### a. Support from others

Carers who used AT for moving and handling of a person with dementia needed ongoing support from formal carers. However, this did not always work as intended.

> “I know Social Services do not allow the [electric] hoist to be used by a single person. Well, all the carers that I know we all did it on our own because there wasn’t anybody else”. [Participant 13]

Even when AT supported a carer in caring tasks, they continued to rely on formal/paid carers to provide much needed respite from constantly caring for a person with dementia.

> “We managed to get a [paid] carer to come in two days a week for three hours so I had a little bit of relief…so it’s had a dramatic effect on my life”. [Participant 4]

In some instances, AT provided access to healthcare professionals, however carers had to provide extra support and time for the person with dementia to access these services.

> “Although, most of it [GP consultation] is done over the phone now. And of course, you know, there was one, there’s been several calls where you had to log into a website…in order to have the [virtual] face to face consultation, which has been really complicated…there’s no way she would have done that on her own”. [Participant 15]

#### b. Ethical issues

Carers clarified ethical issues associated with consent and continued use of AT when cognitive and behaviour issues worsened. Carers let persons with dementia assume AT were innocuous devices and this encouraged the continued use of the AT.

> “At the moment she thinks that it [GPS tracker] controls her house-key. She doesn’t realise it’s a tracker. So, she never ever leaves it because she knows she can’t get back in the house”. [Participant 14]

Carers did set up what could be seen as intrusive AT with consent from a person with dementia, but continue to use the AT, even when ongoing consent may be an issue, due to the progressive cognitive decline.

> “So we’ve got, we set up internal cameras with her full knowledge and agreement but I don’t think she remembered about those after a while”. [Participant 2]

Carers also were aware of and keen to avoid privacy and data leaking issues, when sensitive information could go out externally.

> “Those smart speakers are a brilliant idea in theory, but I feel very unhappy about the idea that every single sound that goes into that thing [smart home system] goes back in some form or another to whoever”. [Participant 20]

## DISCUSSION

The prevalence of dementia is steadily increasing[26]. The attention on person-centred care has enabled people with dementia to live longer lives in the community. Without adequate support for their progressive physical and cognitive needs, the majority of persons with dementia may end up in care homes. Predictions are that availability and ability of carers to continue to support persons with dementia in the community will not match demand[27]. Using technology and especially AT is seen as a solution in supporting persons with dementia and carers. This study explains and describes the impact of AT use among carers who look after persons with dementia living at home. Carers view AT as an adjunct to providing care for the person with dementia and in addition use AT themselves for leisure and improved social contacts and mental wellbeing. Carers continued to modify existing ‘off the shelf’ AT to meet their unique needs and requirements and in most instances, carers were satisfied with the use of AT. We have also explained how AT use in dementia care may not solely rely on the dementia related needs of a person with dementia; it is also dependent on the co-morbidities of persons with dementia and the carers themselves. Carers with existing physical and psychological challenges exacerbated by their own age and health, experience additional burden while caring for a person with dementia and AT use in these instances helps reduce their burden and improves their perceived quality of life. Similar to earlier studies[28] we found that older carers are less likely to access and use new AT, as they may be unaware of these devices and are less likely to request support in acquiring and continued use of these AT. We also explained how carers expressed better satisfaction with AT, when it is integrated into daily life and how they adapted their role and routines to better use the AT in supporting the person with dementia at home. Dementia changes the dynamics of the relationships between the person with dementia and their carer[29], while caregiving may be perceived as stressful, we found that AT does tend to strengthen the caregiving relationship, as the AT itself becomes a socially connective device and facilitates additional time spent using the AT together, especially for leisure and communication. While there exists sophisticated AT and smart homes, we found the use of this alongside simpler devices and non-electronic AT enhanced satisfaction and met carers needs, as reiterated in one of the largest AT Trial[30]. This study also highlights that AT augments the amount of time carers provide care for a person with dementia, however there is a need for respite care[31,32] for carers, and social communication with wider family members who could look after the person with dementia and substitute for the main carer cannot be replaced with AT solutions. Most carers did not consider data sharing and privacy issues connected with AT use, however some carers were specifically concerned about this and limited the type of AT being used, this is likely to require further research and policy intervention as AT devices become sophisticated and ubiquitous in use. This study reinforces earlier findings[4,18] of a mixed picture of use and usefulness of AT and adds to the literature on the impact of AT use on carers. AT alone or in combination with simple devices, formal carers and other interventions has the potential to be an important addition to efforts to improve the safety and welfare of people with dementia, who wish to continue to live at home and that of carers who support them.

### Strength and Limitations

The interviews provided an opportunity to explain and delve deeper into the findings of the survey that explored the experiences and impact of AT use in dementia care amongst carers. Selecting participants from across the UK ensured that geographical limitations of procuring and using AT did not limit the transferability of our findings. The interviews were conducted over telephone due to restrictions from the COVID-19 pandemic and this might have missed out on non-verbal cues and interactions. Despite our repeated efforts, only one participant in the interview, was from an ethnic minority group. It would be important to further investigate how carers from Black, Asian or other Minority Ethnic communities in the UK are using AT and if they are impacted by using AT.

## CONCLUSIONS

This study demonstrates that AT has a beneficial impact on carers who look after persons with dementia. AT devices with the right functionality and when used at those times when needs arose are regarded as useful and satisfactory and are integrated into daily life and routines. Technology development for dementia care requires involvement and consultation with carers. There is a requirement for policy, funding and clinical practice change to a long-term care model that not only facilitates prevention and risk reduction but also encourages education and accessibility of AT to all carers.

## Supporting information

Supplemental file 1_participant information sheet

supplemental file 2_consent form

## Data Availability

The datasets generated during the study are available from the corresponding author on reasonable request.

## Abbreviations

AT: Assistive Technology
CATEQ: Carers Assistive Technology Experience Questionnaire
SF-12: 12-item Short Form Health Survey (version 1)

## Declarations

### Ethics approval and consent to participate

*This study was granted ethical approval by the University of Oxford Central University Research Ethics Committee (Reference number: R57703/RE001). Participants gave informed consent for the survey and interviews*.

### Consent to publish

*Not required*

### Availability of data and materials

*The datasets generated during the study are available from the corresponding author on reasonable request*.

### Authors’ contributions

*VS, CJ and MP conceived the design of the study. VS drafted this version of the manuscript with critical revision and input from MP and CJ. All authors have read and given approval for this version. VS is the guarantor of the manuscript*.

### Authors’ information

*VS is an Occupational Therapist and postgraduate student registered for his DPhil at the University of Oxford exploring informal carers’ experience of assistive technology use in dementia. MP is an Associate Professor within the Health Services Research Unit (HSRU), Nuffield Department of Population Health, University of Oxford. CJ is Professor of Health Services Research and Director of the HSRU, Nuffield Department of Population Health, University of Oxford. MP and CJ have extensive experience in research methods and are joint supervisors of VS for the DPhil*.

## Acknowledgements

*Authors would like to acknowledge support from the three members of the patient and public engagement and involvement group set up as part of the carers’ experience of assistive technology use in dementia study*.

