## Supplemental file 1_participant information sheet for "Carers using assistive technology in dementia care: an explanatory sequential mixed methods study"

Nuffield Department of
POPULATION HEALTH
Medical Sciences Division

UNIVERSITY OF OXFORD

Professor Crispin Jenkinson

Telephone number: 01865 289441
Dr Michele Peters

Telephone number: 01865 289428
Vimal Sriram (Doctoral Student)

Telephone number: 01865 743762
Health Services Research Unit, Nuffield Department of Population Health
Richard Doll Building, Old Road Campus, Oxford OX3 7LF

### Carers' experience of Assistive Technology (AT) use in dementia
#### PARTICIPANT INFORMATION SHEET

Ethics Approval Reference: R57703/RE001

##### 1. *What is the purpose of this study?*

Dementia describes a set of symptoms that may include memory loss and difficulties with thinking, problem solving or language. Caring for a person with dementia can be demanding for informal carers (family, friends and neighbours) and can affect their mental and physical health and their social lives.  Assistive Technology (AT) devices are often electronic. They include talking clocks, electronic medication dispensers, smart gas meters, falls and motion detectors and door exit alarms. While AT is usually aimed at helping the person with dementia, these may also have an impact on carers. Due to the thinking and problem-solving difficulties the person with dementia may have, the carer may need to be an active user of the AT. It is not yet clear what positive or negative effects such technology may have on carers and there is little information on their experience with its use.

<u>Purpose of this research:</u>
This research will explore the experiences of Assistive Technology from the perspective of those looking after and helping persons with dementia at home. It will investigate the types of AT used and preferred, the purposes, benefits and disadvantages of the AT, and how the AT impacts on their burden and wellbeing.

##### 2. *Why have I been invited to take part?*

You have been invited because you participated in a survey of the 'Carers' experiences of AT use in dementia project' and indicated, that we can invite you for an interview.
To participate in the interview, you need to be:

- Continuing to provide help or support to a person with dementia (e.g. shopping, leisure, personal care, finance).

##### 3. *Do I have to take part?*

No, your participation is voluntary.  You can ask questions about the study before deciding whether to take part. If you agree to take part, you may withdraw from the study at any time, without any penalty and without giving a reason. If you choose to withdraw after the interview, the research team will delete any data including personal information and interview recordings and transcripts, and it will not be used in the analysis.

4. ***What will happen if I take part in the study?***
If you are happy to take part, you will be asked to answer questions in an informal interview, like a conversation. The interview questions will ask you about the impact and effects of using Assistive Technology in helping care for your relative/friend/neighbour. The interview will be audio recorded to allow us to type up your answers. You will never be identified by any of your personal information.
The interview should take approximately 60-90 minutes and will take place at your home, your place of work, by telephone or at the University of Oxford. The interview location and time will be arranged in discussion with you, to suit your convenience and preference. The interviews will be conducted by Mr Vimal Sriram, a doctoral student at the University of Oxford.

5. ***Are there any potential risks in taking part?***
The questions asked during the interview may be personal and occasionally some people feel upset when asked to think about their experiences of looking after a person with dementia. You do not have to answer any question that you would prefer not to answer. If you become upset at any point, the researcher will ask you if you wish to pause or stop the interview. You could then: stop and withdraw your data (the interview recording would be deleted), end the interview and allow the interview recording until that point to be used in the research, or carry on with the interview when you are ready.
The researcher can provide you with an information sheet which contains a list of organisations who you can get in touch with if you feel the need for further support.

6. ***Are there any benefits in taking part?***
You will not receive any direct benefit by taking part in this study. However, the information gained in this research study will provide a better understanding and insight of carers' experiences of using AT and its impact on their well-being. This can be used by health professionals and researchers to support other carers of persons with dementia who use AT.

7. ***What happens to my data?***
The **research data** will be stored and examined using University approved software.
Any information that you have given in the interview that could identify you will be removed from the interview before it is analysed. Confidentiality will be maintained throughout this research study. If you consent to take part, you will be required to sign an informed consent form. To protect your identity, your name will be replaced by a pseudonym in any research reports. Any identifying information like your name, details or other personal information will not be used or disclosed to anyone outside, to any third party or appear on any transcripts, thesis, publications or on any academic paper.
However, there might be certain circumstances in which it may be necessary to breach this confidentiality and disclose information to a third party. This includes situations when someone provides information during the study that raises serious concern about:

- Intention to harm themselves or other people
- Risk to the health, welfare or safety of vulnerable adults such as someone with dementia

- Disclosure of a criminal offence

The researcher will discuss this issue with you before telling anyone else. The researcher will be obliged to share this evidence with his supervisor, who may advise that further action is taken.

Personal / sensitive information such as your name, age, gender, marital status, employment status, telephone number or address details in case of face-face interviews will be stored confidentially using computer software that does not allow anyone else except the researcher and his supervisors access to your data. All paper forms will be stored in a locked cupboard within the Department of Population Health, University of Oxford. Your personal/sensitive data, including your signed consent forms will be kept separately from audio recordings and transcripts from your interviews. Your answers may be quoted directly in the research publication with information suitably anonymised. All audio recordings will be erased permanently once they have been transcribed. All research data and records will be stored for a minimum retention period of 3 years after publication or public release of the work of the research.

8. ***Will the research be published?***

The research will be written up as a doctoral thesis. On successful submission of the thesis, it will be deposited both in print and online in the University archives, to facilitate its use in future research. The thesis will be published open access.

Additionally, the research may be published in academic journals and presented in national and international conferences. The University of Oxford is committed to the dissemination of its research for the benefit of society and the economy and, in support of this commitment, has established an online archive of research materials. This archive includes digital copies of student theses successfully submitted as part of a University of Oxford postgraduate degree programme. Holding the archive online gives easy access for researchers to the full text of freely available theses, thereby increasing the likely impact and use of that research.

9. ***Who is organising and funding the research?***

This study is carried out as part of the DPhil (PhD) Programme in Population Health at the Nuffield Department of Population Health, University of Oxford.

10. ***Who has reviewed this study?***

This study has been reviewed by, and received ethics clearance through, the University of Oxford Central University Research Ethics Committee (Reference number: R57703/RE001).

11. ***Data Protection:***

The University of Oxford is the data controller with respect to your personal data, and as such will determine how your personal data is used in the study. The University will process your personal data for the purpose of the research outlined above. Research is a task that we perform in the public interest. Further information about your rights with respect to your personal data is available from http://www.admin.ox.ac.uk/councilsec/compliance/gdpr/individualrights/."

12. ***Who do I contact if I have a concern about the study or I wish to complain?***

If you have a concern about any aspect of this study, you can contact the researcher through an email at or by telephone on 01865 743762 or my supervisors Dr Michele Peters or by telephone on 01865 289428 or Professor Crispin Jenkinson or by telephone on 01865 289441, who will do their best to answer your query. We will acknowledge your concern within 10 working days and give you an indication of how we intend to deal with it. If you remain unhappy or wish to make a formal complaint, please contact the chair of the Research Ethics Committee at the University of Oxford who will seek to resolve the matter in a reasonably expeditious manner:
Chair, **Medical Sciences Inter-Divisional Research Ethics Committee**;; Address: Research Services, University of Oxford, Wellington Square, Oxford OX1 2JD

**13.** ***Further Information and Contact Details***

The interviews for this researcher study will be carried out by Mr. Vimal Sriram (Doctoral student) from the Nuffield Department of Population Health, University of Oxford. The researcher will identify himself to you using a University of Oxford student card.

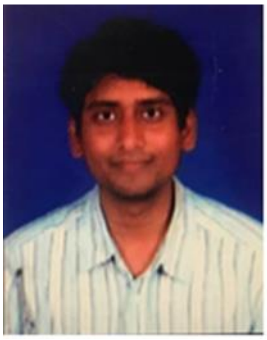

If you would like to discuss the research with someone beforehand (or if you have questions afterwards), please contact:

Mr. Vimal Sriram
Nuffield Department of Population Health
Health Services Research Unit
Richard Doll Building, Old Road Campus, Oxford OX3 7LF
Telephone number: 01865 743762


**Thank you for taking the time to read this information sheet.**
