## supplemental file 2_consent form for "Carers using assistive technology in dementia care: an explanatory sequential mixed methods study"

Nuffield Department of
POPULATION HEALTH
Medical Sciences Division

UNIVERSITY OF OXFORD

Professor Crispin Jenkinson

Telephone number: 01865 289441
Dr Michele Peters

Telephone number: 01865 289428
Vimal Sriram (Doctoral Student)

Telephone number: 01865 743762
Health Services Research Unit
Nuffield Department of Population Health
Richard Doll Building, Old Road Campus, Oxford OX3 7LF

### PARTICIPANT CONSENT FORM

CUREC Approval Reference: R57703/RE001

#### Carers' Experience of Assistive Technology (AT) use in dementia

Purpose of Study:
This research explores the experiences of Assistive Technology use from the perspective of those looking after or helping persons with dementia. The research will look at types of Assistive Technology used and its benefits and dis-advantages. We will explore facilitators or barriers to using Assistive Technology and its impact on burden and well-being.

| | | *Please initial/tick each box* |
|---|---|---|
| 1 | I confirm that I have read and understand the information sheet version 0.2 dated June 2018 for the above study. I have had the opportunity to consider the information, ask questions and have had these answered satisfactorily. | |
| 2 | I understand that my participation is voluntary and that I am free to withdraw at any time, without giving any reason, and without any adverse consequences or academic penalty. | |
| 3 | I am over 18 years of age | |
| 4 | I understand that research data collected during the study may be looked at by designated individuals from the University of Oxford where it is relevant to my taking part in this study. I give permission for these individuals to access my data. | |
| 5 | I understand that this project has been reviewed by, and received ethics clearance through, the University of Oxford Central University Research Ethics Committee. | |
| 6 | I understand only the researchers directly involved in this study will have access to personal data provided, how the data will be stored and what will happen to the data at the end of the project. | |

Professor Crispin Jenkinson

Telephone number: 01865 289441
Dr Michele Peters

Telephone number: 01865 289428
Vimal Sriram (Doctoral Student)

Telephone number: 01865 743762
Health Services Research Unit
Nuffield Department of Population Health
Richard Doll Building, Old Road Campus, Oxford OX3 7LF

| | | |
|---|---|---|
| 7 | I Understand there might be certain circumstances (as outlined in section 7 of the participant information sheet) in which it may be necessary to breach this confidentiality and disclose information to a third party | ☐ |
| 8 | I understand how this research will be written up and published. | ☐ |
| 9 | I understand how to raise a concern or make a complaint. | ☐ |
| 10 | I consent to the interview being audio recorded | ☐ |
| 11 | I understand the audio recordings will only be used to transcribe the interview accurately and will be anonymised in research outputs | ☐ |
| 12 | I give permission to be quoted directly in the research publication suitably anonymised | ☐ |
| 13 | I agree to take part in the study | ☐ |

| | | |
|---|---|---|
| Name of Participant | Date | Signature |
| Name of person taking consent | Date | Signature |
